# Meal Effects on Gastric Bioelectrical Activity Utilizing Body Surface Gastric Mapping in Healthy Subjects

**DOI:** 10.1101/2023.10.31.23296947

**Authors:** I-Hsuan Huang, Stefan Calder, Armen A. Gharibans, Gabriel Schamberg, Chris Varghese, Christopher N. Andrews, Jan Tack, Greg O’Grady

## Abstract

**Background:** Gastric sensorimotor disorders are prevalent. While gastric emptying measurements are commonly used, they may not fully capture the underlying pathophysiology. Body surface gastric mapping (BSGM) recently emerged to assess gastric sensorimotor dysfunction. This study assessed varying meal size on BSGM responses to inform test use in a wider variety of contexts.

**Methods:** Data from multiple healthy cohorts receiving BSGM were pooled, using four different test meals. A standard BSGM protocol was employed: 30-min fasting, 4-hr post-prandial, using Gastric Alimetry® (Alimetry, New Zealand). Meals comprised: i) nutrient drink + oatmeal bar (482 kcal; ’standard meal’); ii) oatmeal bar alone; egg and toast meal, and pancake (all ∼250 kcal). Gastric Alimetry metrics included BMI-adjusted Amplitude, Principal Gastric Frequency, Gastric Alimetry Rhythm Index (GA-RI) and Fed:Fasted Amplitude Ratio (ff-AR).

**Key Results:** 238 participants (59.2% female) were included. All meals significantly increased amplitude and frequency during the first post-prandial hour (p<0.05). There were no differences in postprandial frequency across meals (p>0.05). The amplitude and GA-RI of the standard meal (n=110) were significantly higher than the energy bar alone (n=45) and egg meal (n=65) (all p<0.05). All BSGM metrics were comparable across the 3 smaller meals (p>0.05). A higher symptom burden was found in the oatmeal bar group vs the standard meal and pancake meal (p=0.01, 0.003, respectively).

**Conclusions & Inferences:** The consumption of lower calorie meals elicited different post-prandial responses, when compared to the standard Gastric Alimetry meal. These data will guide interpretations of BSGM when applied with lower calorie meals.

## INTRODUCTION

Gastric sensorimotor disorders are prevalent in the general population ^1–3^. Gastric motility dysfunction plays a pivotal role in explaining symptoms and as a potential therapeutic target in patients with not only motility disorders but also in disorders of gut–brain interaction ^4^. While gastric emptying measurement is a widely used test for evaluating upper gastrointestinal symptoms, the pathophysiology of clinical symptoms is insufficiently captured by the gastric emptying test alone in this heterogenous population ^5^ ^6^. Recently, along with advances in understanding of the electrophysiological basis of stomach, high-quality gastric myoelectrical activity (GMA) recordings to assess neuromuscular function noninvasively using a provocative test meal have become available in the clinical setting ^7^. This high-resolution electrical mapping technique, also known as body surface gastric mapping (BSGM), enables accurate cutaneous gastric bioelectrical recording and provides valuable insights into the pathophysiology of gastric dysmotility in addition to gastric emptying measurements ^5,8^.

The first commercially available BSGM test, the Gastric Alimetry™ System (Alimetry, New Zealand), has undergone rigorous analytical and clinical validation ^5,9–12^. A standardized test meal consisting of a nutrient drink and oatmeal energy bar (482kcal) was suggested in the protocol and appropriate normative references have been established ^13^. However, the optimal test meal for symptomatic individuals is uncertain, as some patients may have difficulty tolerating the relatively large meal ^8^. In addition, smaller meals may be desirable to evaluate patients after gastric operations or in other contexts ^14^.

In Europe, gastric emptying breath tests are commonly used for measuring gastric emptying rates due to non-invasiveness and absence of radioactive labeling. These tests typically employ test meals that have been widely utilized in clinical practice and are generally well-tolerated ^15^. Therefore, employing the same test meal for BSGM as used in the emptying test could augment the BSGM procedure and facilitate its integration into the emptying breath test as a multimodal assessment. Previous studies using legacy electrogastrography (EGG) tests reported that the consumption of reduced-calorie meals did not evoke significant changes in metrics from the preprandial to the postprandial period ^16,17^. However, whether a lower-calorie test meal, as used in routine gastric emptying breath tests, can be an alternative test meal option and elicit the expected postprandial physiological responses in high-resolution BSGM has not been systematically addressed.

Hence, the aims of this study were to examine the impact of different meals on BSGM and postprandial symptom profiles in healthy individuals. These data will be valuable in order to establish meal validity and inform whether an adjusted normative reference range is necessary for the use of BSGM with the emptying breath test meal and in other research contexts where a lower-calorie stimulus is desired.

## METHODS

This study was conducted with multiple cohorts of healthy subjects receiving four different types of meal during BSGM, pooled from a database of diverse control participants in Auckland, New Zealand, Calgary, Canada, Leuven, Belgium, Louisville, Kentucky, United States and Western Sydney, Australia. Ethical approval was obtained (ethical approval references: Auckland Health Research Ethics Committee (AHREC) - AH1130, Conjoint Health Research Ethics Board (CHREB) - REB19-1925, The Ethics Committee Research UZ Leuven S65541, The University of Louisville Institutional Review Board - 723369, and Western Sydney University Human Research Ethics Committee - H13541). All participants provided written informed consent.

### Participants

Healthy volunteers were recruited via local advertisement and e-mail. They were included if they were aged above 18 years, without history of abdominal surgery and taking any medication known to affecting gastrointestinal function. Subjects meeting diagnostic criteria of disorders of gut brain interactions according to Rome IV Criteria were excluded. Specific exclusion criteria related to BSGM, per the Gastric Alimetry Instructions For Use (IFU) were BMI >35 kg/m^2^, active abdominal wounds or abrasions, fragile skin, and allergies to adhesives.

### Study Protocol for BSGM

BSGM was performed using the Gastric Alimetry System (Alimetry, New Zealand), employing stretchable electronics attached to a wearable Reader placed over the epigastrium (8x8 electrodes; 225 cm^2^) ^8,18^. The standard Gastric Alimetry test protocol was followed, as described in previous literature ^19^. All studies were started in the morning, with participants fasting for >6 hrs and asked to avoid caffeine and nicotine on the day prior to testing. Array placement was preceded by shaving if necessary, skin preparation (NuPrep; Weaver & Co, CO, USA), and customized placement according to patient surface anatomy measurements ^18^. Recordings were performed for a fasting period of 30 minutes, followed the specified test meal consumed within 10 minutes and a 4-hr postprandial recording in order to capture a full gastric activity cycle. Participants sat reclined in a chair and were asked to limit movement, talking, and sleeping, but were able to read, watch media, work on a mobile device, and mobilize for comfort breaks. The subjects rated early satiation after meal completion. Symptoms of nausea, bloating, upper gut pain, heartburn, stomach burn, and excessive fullness were measured during the tests via the validated Gastric Alimetry App at 15-minute intervals using 0-10 visual analog scales (0 indicating no symptoms; 10 indicating the worst imaginable extent of symptoms) and combined to form a “Total Symptom Burden” score ^20^.

### Test meals

Following a 30-minute fasting period BSGM recording, four types of test meals accessible in the clinical setting were administered. The standard Gastric Alimetry test protocol involved a larger meal (482 kcal) consisting of a common supplemental nutrition drink (Ensure, Abbott Nutrition, IL, USA, 232 kcal, 250 mL, 9 g fat, 34 g carbohydrate, 9 g protein) along with an oatmeal energy bar (Clif Bar & Company, CA, USA, 250 kcal, 5 g fat, 45 g carbohydrate, 10 g protein, 7 g fiber) ^21^. In addition, three commonly used and easily prepared smaller meals, each providing approximately 250 kcal, were given. These included an oatmeal energy bar with 250ml of water (Clif Bar; see above), a pancake with 180 ml water (11.2 g fat, 31.7 g carbohydrate, 8.4 g protein) ^22^, and an egg with two slices of white toast and 180 ml of water (9.4 g fat, 34 g carbohydrate, 11.8 g protein)^23^. The latter two meals have also been applied as part of the routine gastric emptying breath test ^24^.

### Data Analysis

The standard metrics derived from the BSGM test were analyzed for the healthy subjects in each group ^25^. BSGM raw signal processing included the standard Gastric Alimetry filtering, artifact detection, and signal recovery techniques ^26^.

The spectral analysis included BMI-adjusted Amplitude, Principal Gastric Frequency, Gastric Alimetry Rhythm Index™ (GA-RI)™ and Fed:Fasted Amplitude Ratio (ff-AR). Amplitude (μV) reflects combined electrical response activity and electrical control activity of the stomach ^27^. BMI correction was applied to achieve a BMI-adjusted reference interval using a multiplicative regression model ^25^. Principal Gastric Frequency (cycles per minute, cpm) is the measurement of the sustained frequency within the plausible gastric range to avoid influence from motion artifacts and periodic adjacent colonic activity ^28^. GA-RI is a measure of concentrated gastric activity within a narrow gastric frequency band over time relative to the residual spectrum. The GA-RI is scaled between 0 (no rhythm stability) and 1 (maximum rhythm stability) and is independent of frequency, with a normative reference range cutoff of 0.25. The revised ff-AR metric measures the gastric response based on the maximum amplitude in any single 1-hour period of a 4-hour postprandial window.

### Statistical analysis

Quantitative data are presented as the median and interquartile range (IQR 25–75) for variables not normally distributed. For continuous variables, Mann–Whitney U test was applied for two groups comparison. Pre- and post-prandial parameters from BSGM were compared using a Wilcoxon signed-rank test. For the analysis of multi-group comparisons, the Kruskal–Wallis test was performed. Once a significant difference between the four groups had been determined, pairwise comparison of groups was applied with the Dunn-Bonferroni test. Pearson’s chi-square test or Fisher’s exact test was performed for categorical variables. Throughout, the statistical significance level used was p<0.05. All statistics have been performed with SPSS 22 (SPSS) for Windows.

## RESULTS

### Study subjects

A total of 238 study participants (59.2% female, median age 29 (23-38) years, and BMI 22.7 (21.2-25.6) kg/m2) were included. Out of all participants, 110 subjects received the larger standard meal with an energy bar and nutritional drink. 45 participants received an energy bar along with water, while 65 participants received an egg meal, and 18 participants received a pancake with water. The percentage of 100% meal completion was significantly higher with a smaller meal (97.7% vs 89.1%, p<0.01). The baseline characteristics of subjects in 4 groups are shown in Table 1.

**Table 1.** Baseline characteristics of subjects with four kinds of meals.

|  | **Bar + Ensure**<br>**(N=110)** | **Bar + Water**<br>**(N=45)** | **Egg meal**<br>**(N=65)** | **Pancake**<br>**(N=18)** | **P**<br>**value** |
| --- | --- | --- | --- | --- | --- |
| **Female** | 55.5% | 77.8% | 50.8% | 66.7% | 0.04 |
| **Age (years)** | 32 (24-51) | 27 (22-34) | 29 (23.5-34) | 26.5 (23.8-32.3) | 0.04 |
| **BMI (kg/m<sup>2</sup>)** | 23.8 (21.4-26.9) | 23.4 (21.6-25.5) | 22.1 (21.0-24.2) | 22.4 (20.2-24.3) | 0.06 |
| **Percentage successfully**<br>**completing 100% of meal** | 89.1% | 93.3% | 100% | 100% | 0.01 |
| **Mean meal completion** | 97,1%±10.3 | 99.3%±2.5 | 100% | 100% | 0.02 |

### Effects of test meals on BSGM metrics

Summary spectral plots and symptom burden graphs for each test meal are depicted in Figure 1. All meals showed a distinct marked post-prandial deviation in amplitude, and minor deviation in frequency. Comparing to three 250-kcal meal, the amplitude curve corresponding to the larger meal (Bar + Ensure) was characterized by a more extended postprandial response before returning to baseline amplitude levels. Quantitative analyses of the test meals for the overall recording period are shown in Figure 2.

**Figure 1.**
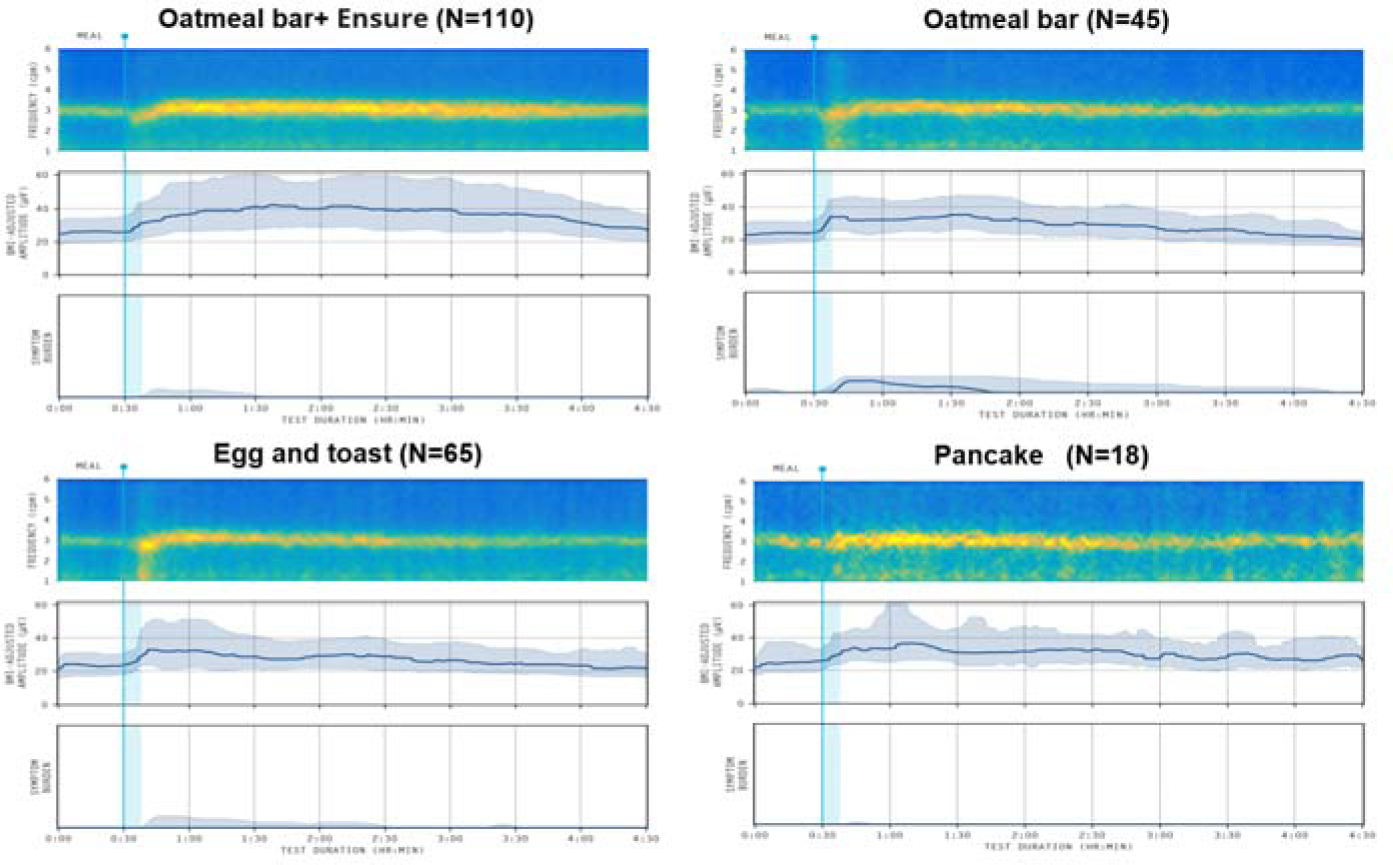
Summary of spectral phenotypes arising from BSGM and symptom burden in each different meal, median (IQR shaded)

**Figure 2.**
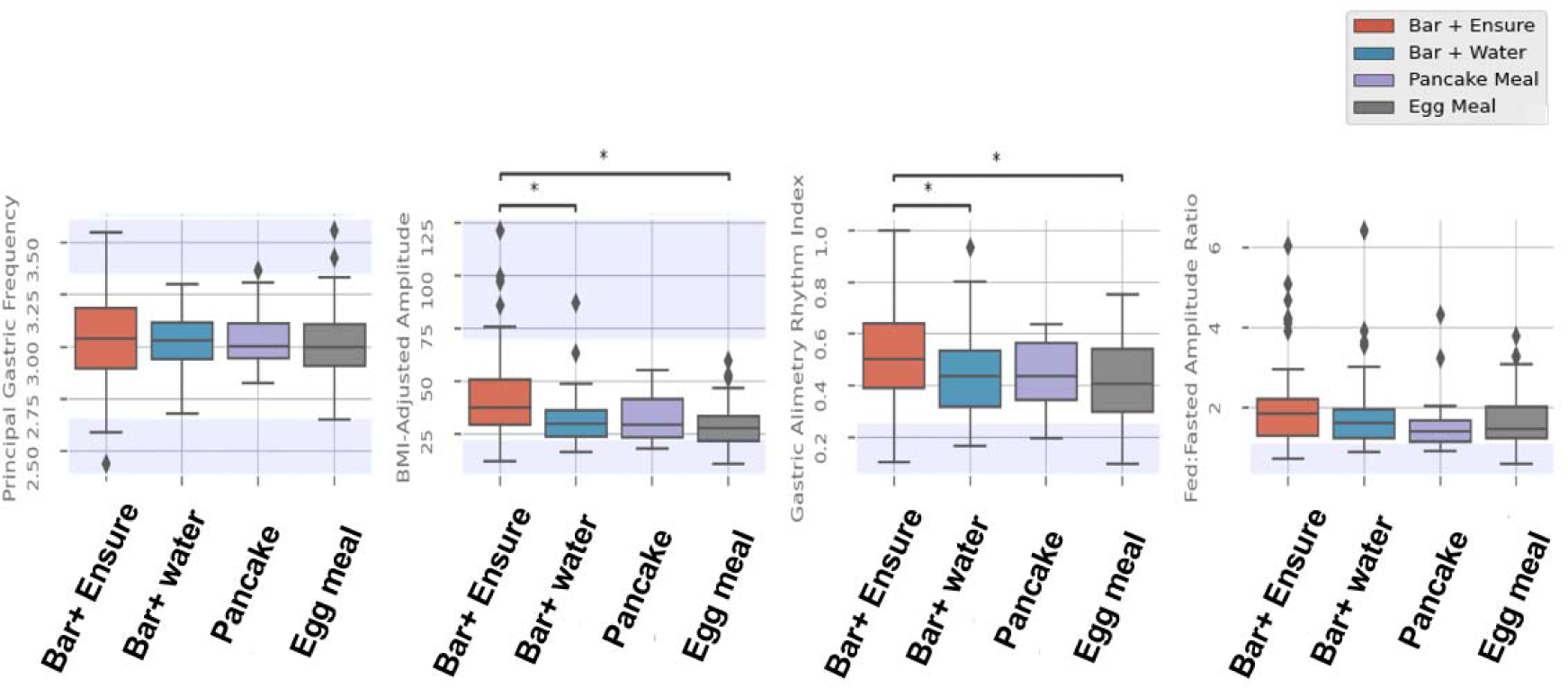
Comparative analysis of BSGM metrics during overall recording period across different meal types. * p<0.05.

There were no significant differences in overall frequency among the four meals (p=0.853). The averaged BMI-adjusted amplitude and GA-RI of the standard meal was significantly higher than those of energy bar alone and egg meal (amplitude p=0.005, <0.001; GA-RI p=0.048, 0.001, respectively). However, when compared to the pancake, the differences did not reach statistical significance (p=0.167, 0.830, respectively), while noting the smaller sample size in this latter meal group. There was no significant difference in amplitude and GA-RI between the three smaller meals (all p>0.05). The ff-AR also showed no significant difference in terms of pairwise comparisons between individual small meals and the standard test meal (energy bar, pancake, egg-based meal to standard meal, p=0.390, 0.122, 0.096, respectively).

Similar results were observed within the average across entire 4-hour postprandial period. There was no difference in frequency between each meal (all p>0.05). The average amplitude and GA-RI of the standard meal were significantly higher than those of the energy bar alone and the egg meal (amplitude p=0.003, <0.001, respectively; GA-RI p=0.037, 0.002, respectively). However, it did not achieve significance when comparing to the pancake group (p=0.130, 0.655, respectively). Notably, there were no significant differences in postprandial amplitude and GA-RI among the three smaller meals (all p>0.05).

Figure 3 presents a comparative analysis of three metrics for each meal across hourly intervals. Throughout each hour, there were minimal variations in frequency between the meals. However, the average amplitude and GA-RI of the standard meal were significantly higher than those of the smaller meals. This distinction became more pronounced as time advanced beyond the initial postprandial hour, suggesting a lasting effect of the larger meal on GMA.

**Figure 3.**
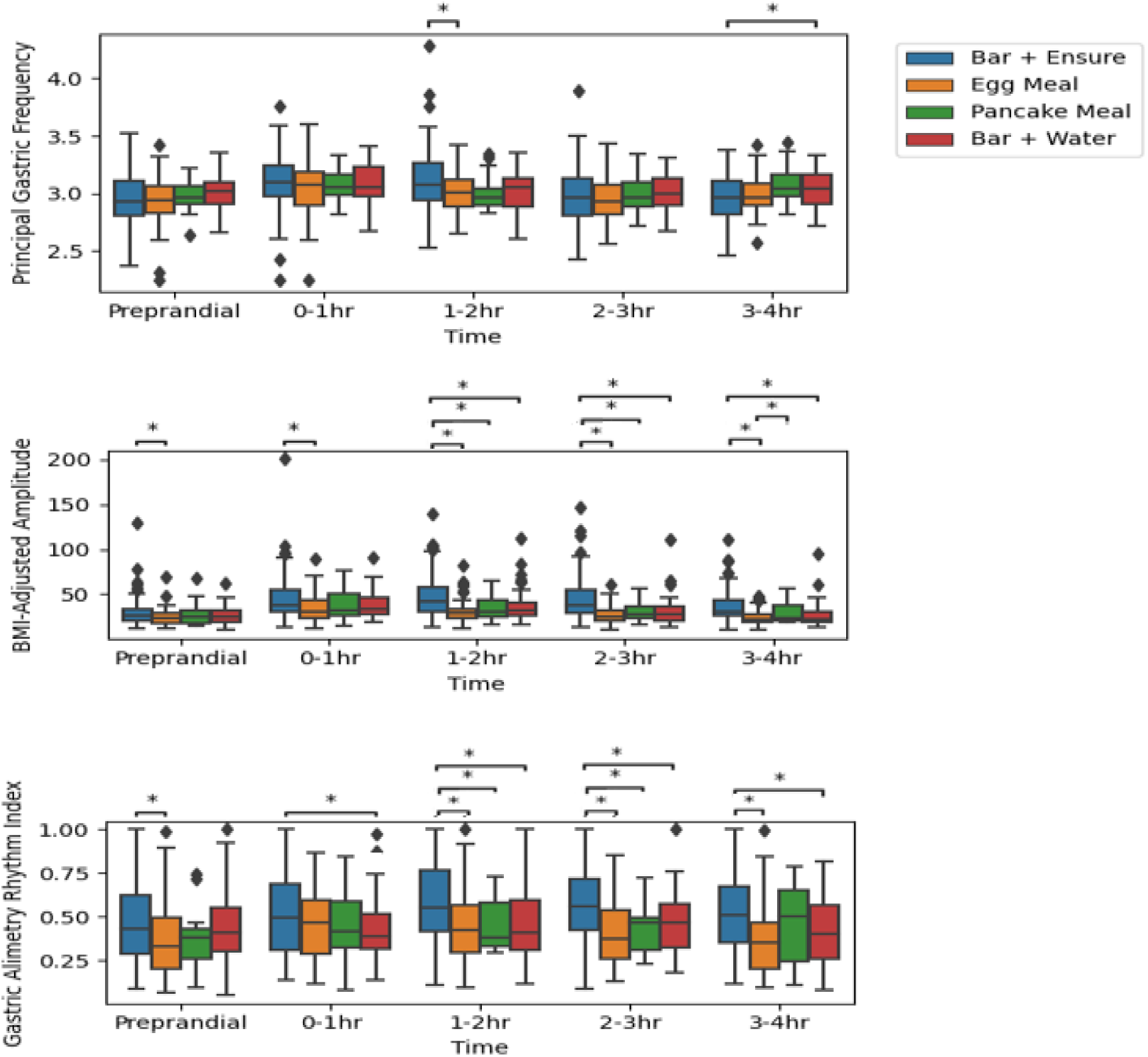
Comparison of the frequency, amplitude, and rhythm stability for each meal at every hour. * p<0.05.

All meals led to a significant post-prandial increase in Principal Gastric Frequency and BMI-adjusted amplitude during the first hour after consumption (Table 2). The post-prandial frequency showed a range of increases from 0.135 to 0.100 cpm across the different meal groups. There was no significant difference among the four meals (p=0.849). The increases in amplitude exhibited more variability, ranging from 49.8% to 17.4%, with the energy bar meal resulting in the largest increase and the pancake meal showing the smallest. However, these differences did not reach statistical significance (p=0.428). Additionally, the GA-RI showed a significant increase after consuming the standard meal and egg meal, while there was no significant change observed after consuming the oatmeal bar and pancake (Table 2).

**Table 2.** Effect of meals on BSGM parameters, median (25th to 75th percentile)

|  | **Bar + Ensure**<br>**(N=110)** | **Bar + Water**<br>**(N=45)** | **Egg meal**<br>**(N=65)** | **Pancake**<br>**(N=18)** |
| --- | --- | --- | --- | --- |
| **BMI-adjusted**<br>**amplitude (µV)** |  |  |  |  |
| **Fasting** | 26.6 (20.8-32.9) | 23.9 (18.7-32.5) | 23.3 (17.2-28.8) | 25.2 (17.9-32.4) |
| **First hr Postprandial** | 38.0 (30.3-55.0) | 33.8 (27.5-46.5) | 30.1 (23.3-46.2) | 31.6 (25.4-51.7) |
| **Percentage increase** | 45.3% (10.7-76.3) | 49.8% (11.6-81.5) | 30.6 % (0.0-66.1) | 17.4% (-1.1-63.6) |
| **P value** | <0.001 | <0.001 | <0.001 | 0.021 |
| **Frequency (CPM)** |  |  |  |  |
| **Fasting** | 2.93 (2.80-3.11) | 3.02 (2.91-3.10) | 2.94 (2.82-3.08) | 2.96 (2.87-3.08) |
| **First hr Postprandial** | 3.10 (2.97-3.25) | 3.05 (2.98-3.24) | 3.08 (2.89-3.19) | 3.05 (2.98-3.17) |
| **Frequency change** | 0.135 (0.050-0.265) | 0.105 (0.005-0.295) | 0.108 (0.000-0.276) | 0.100 (0.000-0.903) |
| **P value** | <0.001 | 0.002 | <0.001 | 0.020 |
| **GA-RI** |  |  |  |  |
| **Fasting** | 0.433 (0.288-0.625) | 0.413 (0.297-0.557) | 0.334 (0.198-0.498) | 0.383 (0.178-0.436) |
| **First hr Postprandial** | 0.496 (0.303-0.693) | 0.388 (0.309-0.539) | 0.465 (0.286-0.602) | 0.419(0.318-0.601) |
| **P value** | 0.013 | 0.821 | 0.020 | 0.369 |

### Effects of test meals on symptoms

Minimal postprandial symptom burden was observed after all meal intakes, consistent with the healthy control population studied, with the pancake meal being the most well tolerated option. Meal-related symptoms generally increased at the first hour after the meal and subsequently decreased over time to reach the preprandial values. The oatmeal bar group had significantly higher postprandial Total Symptom Burden Score (0.63 (0.00-2.92)) compared to the other groups (standard meal (0.05 (0.00-0.40)), pancake (0.00 (0.00-0.15)), egg-based meal (0.06 (0.00-0.83))); after pairwise comparison (all p<0.05). The change of total symptom severity between the first post- and pre-prandial hours in the oatmeal bar group was significantly higher than the standard meal and the pancake (p=0.010, 0.003, respectively) but did not reach significance compared to the egg meal (p=0.213). When looking at individual symptoms, scores of ’excessive fullness’ were significantly higher in subjects receiving the energy bar, compared to the other three meals (all p<0.05). Additionally, the energy bar group was associated with a higher severity of nausea than the egg meal (p=0.011). No difference was noted in intensities of bloating, upper gut pain, heartburn and stomach burn between four meals (all p>0.05) (Supplement table 1).

## DISCUSSION

This is the first study comparing the effects of the different meals on gastric function activity utilizing a BSGM device that is capable of defining gastric bioelectrical activity at high-resolution^19^. In legacy EGG literature, an increase in amplitude, frequency and percentage of normal frequency after meal consumption have been described ^17,29^. These changes are considered as part of the meal response in GMA and may reflect the influence of stretch on gastric activity in addition to other co-regulatory factors ^28^. In contrast to legacy EGG studies, which did not show apparent change following a 250-kcal meal ^30^, our results demonstrated a low-calorie meal is able to produce expected postprandial physiological alterations and thus may be recommended for the study of the postprandial response of GMA in symptomatic patients. However, decreasing meal total caloric content decreased the postprandial gastric BMI-adjusted amplitude and GA-RI, while not affecting the frequency or ff-AR.

Our data further quantified the way in which a larger meal invoked a more prolonged meal response, demonstrated by the longer duration prior to the myoelectrical amplitude returning to baseline in comparison to the smaller meals. This observation was expected, and is also consistent with previous gastric emptying studies that reported a longer gastric emptying time with higher calorie-content meals ^30^ ^31,32^. A larger meal may be expected to provoke increased gastric stretch, more vigorous gastric contractions, and greater gastric distension, all of which explain the higher BMI-adjusted amplitude in response to the larger meal. ^33–35^ The higher GA-RI following the larger meal most likely is secondary to the higher amplitude in combination with the more regular and longer-duration period of gastric contractility post-prandially ^25^.

Frequency-based abnormalities in gastric slow waves have been frequently documented in previous electrophysiology studies involving symptomatic patients. As described in the legacy EGG literature, these abnormalities typically involve a reduced duration of normogastria during both fasting and postprandial states, along with an increased occurrence of bradygastria or tachygastria periods ^36,37^. The current study accurately quantifies normal changes in frequency occur following meal ingestion in the healthy human stomach, for the first time using BSGM techniques. These natural fluctuations provide valuable insights into the dynamic nature of GMA during the postprandial period and highlights the potential of postprandial frequency change as a marker of gastrointestinal function.

Our study also found no statistically significant differences were observed in BSGM metrics after consumption of three smaller meals with identical calories. These results align with the findings from a previous randomized trial that controlled several parameters, including caloric intake, fiber content, and meal volume. In this previous trial, reducing the meal volume or increasing the fiber content while keeping the calories consistent did not result in any significant changes in EGG metrics ^17^. However, our study which included robust methods of real-time symptom evaluation did reveal that the higher fiber meal (oatmeal bar) was associated with higher postprandial symptoms.

The availability of a range of test meals is crucial for the wider application of BSGM tests in different clinical scenarios. While the original test meal with a large caloric challenge and mixed liquid/solid meal was selected in order to induce a gastric stressor that would provoke symptoms in diverse gastroduodenal populations ^18,38^, smaller meals may be preferable in patients with lower tolerances, such as nausea and vomiting / gastroparesis patients ^39^, post-surgical populations ^14^, and perhaps paediatric populations in the future ^40^. In a recent study using Gastric Alimetry in nausea and vomiting patients, for example, more than half of patients could not complete the full standard meal (median meal completion 90% (interquartile range 70-100)), although a sensitivity analysis did not show a change in primary outcome with reduced meal consumption ^8^. In addition, simultaneous measurement of gastric bioelectrical activity by legacy EGG with an emptying breath test has been applied in previous studies ^16^. The utilization of a same test meal for routine gastric emptying breath tests offers an opportunity to integrate BSGM into the emptying measurement, allowing for a closer temporal correlation between the two physiological assessments and overcoming challenges related to human resources and time consumption of the test in clinical setting ^5,41^.

To apply BSGM in clinical practice, defining reference intervals in healthy subjects is essential. Current BSGM reference intervals were developed using the larger standard test meal ^38^. Although our results indicated that all meals, including the three lower calorie options, elicited significant post-prandial meal responses, we found some reductions in selected metrics over the post-prandial course, specifically BMI-adjusted amplitude and GA-RI. Therefore, to achieve optimal accuracy in result interpretation, we recommend that the reference ranges ideally should be adjusted when applied to populations when smaller meals are used.

There are several limitations to this study. First, the study is not a crossover design, such that the effect of meals was not measured on the same individuals. Therefore, some covariates in different meal-groups may be unbalanced. Second, the pancake group had a limited number of subjects, which could have influenced the significance of certain variables. However, the consistency across the three smaller meal sizes suggests that the pancake results overall are similar to the comparable egg meal which is also used in breath test emptying studies. Finally, an increase in movement artifacts was observed during the simultaneous administration of the gastric emptying breath test and BSGM. However, to address this issue and to ensure data quality, automatic artifact rejection and correction methods are implemented within the system to minimize the impact of movement-related artifacts and maintain the integrity of the collected data ^10^.

In summary, the consumption of lower calorie meals elicited different post-prandial responses, manifesting in lower postprandial amplitude and rhythm stability when compared to the standard Gastric Alimetry meal used with existing test reference intervals. These findings will be important to correctly interpreting BSGM results when applied in various settings with lower calorie meals.

## Data Availability

All data produced in the present study are available upon reasonable request to the authors

## Supplementary

**Table 1.** Individual symptom severity and total symptom burden after four meals consumption, median (25th to 75th percentile)

|  | **Bar + Ensure**<br>**(N=110)** | **Bar + Water**<br>**(N=45)** | **Egg meal**<br>**(N=65)** | **Pancake**<br>**(N=18)** | **P value** |
| --- | --- | --- | --- | --- | --- |
| **Nausea** | 0.0 (0.0-0.0) | 0.0 (0.0-0.0) | 0.0 (0.0-0.0) | 0.0 (0.0-0.0) | 0.009 |
| **Bloating** | 0.0 (0.0-0.0) | 0.0 (0.0-0.13) | 0.0 (0.0-0.0) | 0.0 (0.0-0.11) | 0.070 |
| **Upper Gut Pain** | 0.0 (0.0-0.0) | 0.0 (0.0-0.0) | 0.0 (0.0-0.0) | 0.0 (0.0-0.0) | 0.425 |
| **Heartburn** | 0.0 (0.0-0.0) | 0.0 (0.0-0.0) | 0.0 (0.0-0.0) | 0.0 (0.0-0.0) | 0.493 |
| **Stomach Burn** | 0.0 (0.0-0.0) | 0.0 (0.0-0.0) | 0.0 (0.0-0.0) | 0.0 (0.0-0.0) | 0.832 |
| **Excessive Fullness** | 0.0 (0.0-0.15) | 0.58 (0.0-1.95) | 0.0 (0.0-0.36) | 0.0 (0.0-0.0) | <0.001 |
| **Total Symptom Burden** | 0.05 (0.0-0.40) | 0.63 (0.0-2.91) | 0.06 (0.0-0.83) | 0.0 (0.0-0.15) | <0.001 |

